# Proteomic and mass spectrometry-based identification of viral proteins in human tissue samples from the Network for Pancreatic Organ Donors with Diabetes

**DOI:** 10.1101/2024.10.24.24315944

**Authors:** Tanya C. Burch, Naomi L. Hitefield, Margaret A. Morris, Alberto Pugliese, Jerry L. Nadler, Julius O. Nyalwidhe, the JDRF nPOD-Virus Group

## Abstract

**Aims/Hypothesis:** Multiple studies associated enterovirus (EV) infections with type 1 diabetes. The Network for Pancreatic Organ Donors with Diabetes (nPOD) obtained samples from organ donors with/without type 1 diabetes and launched the nPOD-Virus Group to examine viral infections in donor tissues, using complementary approaches. To this end, we aimed to identify virus proteins/peptides in disease-stratified tissues using proteomic and liquid chromatography-mass spectrometry.

**Methods:** nPOD provided specimens from four donor groups: donors without diabetes (ND, n=33), with type 1 diabetes (T1D, n=25), with type 2 diabetes (T2D, n=7), and without diabetes expressing type 1 diabetes-associated autoantibodies (AAb+, n=17; preclinical disease). We studied flash-frozen pancreas tissue chunks, embedded tissue slices, and islets obtained via laser capture microdissection (LCM). We isolated and processed proteins from these specimens for liquid chromatography–mass spectrometry analysis. We utilized different instruments including a Q-Exactive Orbitrap Mass spectrometer and an Orbitrap Fusion Lumos Mass Spectrometer to acquire high resolution, high mass accuracy and high sensitivity MS data using different scanning methods. We used data dependent acquisition (DDA), data independent acquisition (DIA), and parallel reaction monitoring (PRM). Generated mass spectra were processed and used in protein database searches for identification, qualitative and quantitative comparative analyses of viral protein expression in tissue samples.

**Results:** Advanced proteomics were applied to pancreata from 82 disease-stratified nPOD donors. These analyses generated >1,000 individual mass spectra data files. We identified enterovirus peptides from different serotypes in 28 donors, including 11 donors with type 1 diabetes. These serotypes included several previously associated with type 1 diabetes. For some donors, identification of virus peptides by discovery proteomics was validated by targeted mass spectrometry and Western blot.

**Conclusions/Interpretation:** For the first time we applied complementary mass spectrometry-based proteomics to detect viral proteins in disease-stratified pancreas samples. Some pancreata, including several from donors with type 1 diabetes, were infected by enteroviruses based on detection of viral proteins; in several instances we identified serotypes, which has been arduous with other methods. We detected both structural and non-structural viral proteins, the latter essential for replication, suggesting that enteroviruses may replicate in pancreas, perhaps at low level, given the absence of acute infection. The complexity of our methodology limited application to large sample sets, and accordingly we did not aim to demonstrate an association with disease; our data complement associative data generated with other approaches by the nPOD-Virus Group, overall supporting a role for enterovirus infections in type 1 diabetes.

**Research in Context:** *What is already known about this subject?:* There are previous studies that examined an association of enterovirus infections with type 1 diabetes by examining pancreas tissue, but those are largely limited to the assessment of a single viral antigen by immunohistochemistry in pancreas tissues obtained at autopsy from recent onset patients, from either an historical archive, or from a small series of biopsies.

*What is the key question?:* To examine viral infections in the pancreas from the largest collection of organ donors and disease duration using unbiased comprehensive proteomic and liquid chromatography-mass spectrometry methods.

*What are the new findings?:* We present robust mass spectrometry based proteomics data that are validated by complementary western blot results identifying multiple virus proteins from different virus serotypes in pancreas from disease stratified organ donors. Overall, the findings support the existence of chronic or recurrent infections in the pancreas of some patients with type 1 diabetes.

*How might this impact on clinical practice in the foreseeable future?:* These results provide strong rationale for advancing current efforts to prevent or mitigate type 1 diabetes by vaccination and/or anti-viral therapies.

## Introduction

Type 1 diabetes is a chronic disease resulting from the autoimmune destruction of pancreatic β-cells, mediated by autoreactive CD4+ and CD8+ T-cells recognizing islet cell autoantigens. The triggers of islet autoimmunity remain elusive but several viruses are considered potential candidates and have reported associations with type 1 diabetes, including enteroviruses (especially Coxsackievirus B strains, [CVB]), rotavirus, mumps virus, and cytomegalovirus [1–11]. Different enterovirus serotypes are recognized based on their genetic properties and antigenicity. The viral genome typically consists of a positive-sense single stranded RNA with about 7,500 nucleotides in a single open reading frame (ORF) that encodes for a polyprotein. At the 5’ end the polyprotein is flanked by a long untranslated region (5’-UTR) and a short 3’-UTR. The 5’-UTR possesses a variable spacer region and highly conserved internal ribosome entry site (IRES). In coxsackie viruses (CVB), the encoded genome polyprotein is processed co- and post-translationally to yield four structural proteins, VP1, VP2, VP3 and VP4 and seven non-structural proteins, 2A-2C and 3A-3D. The order of genes along the genome for CVB from 3’ to 5’ is as follows: The non-structural proteins 3D, 3C, 3B and 3D, then 2C, 2B and 2A, and finally the structural proteins VP1, VP3, VP2 and VP4. The structural proteins form the icosahedral capsid of the virus. The non-structural proteins include proteases that cleave viral polyprotein and specific host proteins. Some of these virus-mediated processes inhibit host cellular activities that help the virus evade host defense mechanisms and promote viral genome replication and morphogenesis [12–13].

Liquid chromatography mass spectrometry-based proteomics can be utilized for global protein qualitative and quantitative analysis which may not be uncovered by genomic analysis alone. In the context of the nPOD-Virus Group collaborative effort, our objective was to investigate the possible presence of viral proteins and protein signatures in disease-stratified pancreas from nPOD donors using proteomic and mass spectrometry technology. Our approach identified virus peptides from different enterovirus serotypes in pancreas tissue from several organ donors, including donors with type 1 diabetes. Thus, proteomics-based LC/MS/MS demonstrated virus infections in human pancreas for virus strains that have been associated with type 1 diabetes in previous studies and in the context of the overall analyses performed by the nPOD-Virus group.

## Materials and Methods

### Materials, Chemicals and Reagents

We obtained BCA protein and micro-BCA protein assay kits, dithiothreitol (DTT), desalting and C18 SPE affinity purification columns from Thermo Fisher Scientific (Rockford, IL, USA). Acetone, acetonitrile, methanol, diethyl ether, water and formic acid were Optima LC/MS grade and were purchased from Thermo Fisher Scientific. Ammonium bicarbonate and iodoacetamide were purchased from Sigma-Aldrich (St. Louis, MO, USA). Sequencing Grade Trypsin and LysC were from Promega Corporation (Madison, WI, USA).

### Study Approval and Human Pancreatic Tissue

Disease-stratified pancreas tissues were obtained from nPOD (https://www.nPOD.org), University of Florida, Gainesville, Florida, USA. The samples were from deceased, de-identified organ donors and were obtained in accordance to stipulated ethical regulations [14–15]. All samples were de-identified and obtained by nPOD through its partnership organ procurement organizations as approved by the University of Florida Institutional Review Board (IRB), after consent for organ donation and research was obtained from family members. The study of de-identified organ donor tissues is considered not to involve human subjects and is exempted from review by the Eastern Virginia Medical School Institutional Review Board (IRB). nPOD provided specimens were from four cohorts: donors without diabetes (ND, n=33), donors with type 1 diabetes (T1D, n=25), donors with type 2 diabetes (T2D, n=7), and donors without diabetes expressing with type 1 diabetes-associated autoantibodies (AAb+, n= 17), representing the preclinical disease stage. **Supplementary Table 1** describes the main demographic and clinical features of the donors examined, representing a subset of the donors that were overall examined by the nPOD-Virus group, based on sample availability. Tissue samples were provided as flash frozen tissue chunks and optimal cutting temperature (OCT) compound embedded tissues slices for direct processing and pancreatic islets obtained via laser capture microdissection (LCM) from pancreas sections.

### Extraction and Processing of Proteins for Mass Spectrometry

The three different types of tissue samples, flash frozen, OCT embedded tissue and LCM sections were processed using optimized methods for each sample type. Flash frozen tissues were extracted and directly processed using the trifluoroethanol method [16–17]. For the OCT embedded tissue, the mounting media was washed with PBS prior to protein extraction using the TFE protocol prior to liquid chromatography mass spectrometry (LC/MS/MS) as previously described [16–17]. In this approach, pancreas tissues were homogenized in lysis buffer comprising of 50% TFE in 50mM ammonium bicarbonate, pH 8.3, and incubated on ice for 30 minutes at 4°C to solubilize protein. The mixture was fully homogenized by probe sonication using 20 second cycles, 5 times on ice. Next, to maximize protein solubilization, the homogenized mixtures were heated at 60°C for 30 minutes, then sonicated again prior to centrifugation for 15 min at 10,000 x g to remove insoluble material. The supernatant comprising of solubilized protein was collected and the concentration determined using the BCA assay.

Fifty micrograms of protein from each sample were heat-denatured at 95 °C for 5 minutes before adding 10 mM DTT and heating again at 95 °C for 5 minutes. The reduced samples were cooled to room temperature before alkylation using 15mM iodoacetamide for 30 minutes. The final concentration of TFE was reduced from 50% to 5% in a final volume of 500 µl using 50mM ammonium bicarbonate before digestion with trypsin at a 20:1 protein-to protease ratio at 37°C for 18 hrs. Trypsinization was stopped by the addition of formic acid before centrifugation at 20,000 x g for 10 minutes. The peptides were desalted by solid phase extraction using C18 columns and eluted with 80% acetonitrile in 0.1% formic acid. Eluted peptides were dried in a SpeedVac and stored at -80°C before further analysis. The tryptic peptides were solubilized in normalized volumes of 0.1% formic acid and their concentrations determined using a NanoDrop spectrophotometer. The peptide concentrations were adjusted to 0.5 µg/µl for all the samples before using 2 µg of each for analysis. Laser captured microdissected islet samples were processed for LC/MS/MS as previously described [18].

In complementary experiments, immunoprecipitation was performed on pancreas protein lysates using the anti-VP1 DAKO Clone 5D8/1 monoclonal antibody, using standard methods. Briefly, Protein A/G beads (Thermo Fisher) were added to concentration normalized samples and rotated overnight at 4°C. Samples were centrifuged at 3,000 rpm for 5 min at 4°C to pellet the beads and the supernatants were discarded. The beads were washed three times with cold 1 x RIPA buffer and suspended in SDS-PAGE loading buffer and heated at 95℃ for 10 min, then subjected to SDS-PAGE separation. The separated proteins were stained with colloidal Coomassie to visualize protein bands. The protein bands were excised and processed for Gel-LC/MS/MS as previously described [19].

To improve virus peptide identification rates and to test possible concordance with VP-1 immunostaining on tissue sections, we included in the analysis pancreas sections that were also selected based on positive VP1 signals by IHC on FFPE tissues, presence of HLA Class I hyper-expression and detection of insulin staining in residual beta cells using fresh frozen OCT embedded tissues, as determined by related efforts within the nPOD-Virus Group. Thirty-micrometer (30 µM) tissue slices adjacent to those that were positive for VP1, insulin, and HLA Class I hyper-expression and corresponding negative controls were cut from OCT embedded tissues. Proteins were isolated from the sections and processed for LC/MS/MS using the TFE/ABC approach. For these experiments, we analyzed 18 new nPOD cases that included 8 ND donors, 3 AAb+ donors, and 7 donors with type 1 diabetes. The demographics and disease phenotypes for these cases are summarized in **Supplementary Table 2** and are also included in **Supplementary Table 1** which includes all the cases in the study.

### Liquid Chromatography Mass Spectrometry Data Acquisition

Most LC-MS/MS analyses were performed on a Q-Exactive Orbitrap mass spectrometer (MS) (Thermo Fisher) and a Tribrid Orbitrap Fusion Lumos MS (Thermo Fisher), coupled on-line to a nanoflow LC system (Easy Nano 1200, Thermo Fisher). For these analyses we utilized data dependent acquisition (DDA) and data independent acquisition (DIA) methods [20–21]. A limited number of analyses were performed on a 5600 Triple TOF MS (Sciex), and on a Q-TRAP 4000 mass spectrometer (Sciex) coupled to an Eksigent nano-LC system (Sciex). For proteomic analysis, tryptic peptides were resolved using at a normalized concentration of 0.5 µg/µl 0.1% formic acid for each sample prior to LC/MS analysis. Four microliters of the reconstituted peptides corresponding to 2 µg of the peptides were delivered to a trap column (Acclaim PepMap 100 C18, dimensions 0.1 × 2 cm) at a flow rate of 10 µl/min for 10 min using 0.1% formic acid. The trapped peptides were washed, equilibrated and transferred to a 50 cm, 75 µM inner diameter Thermo Scientific™ EASY-Spray C18 analytical column. Peptides were fractionated and injected into the MS using a 110-min gradient from 2% to 32% solvent B (0.1% FA, 80% in acetonitrile, ACN) at a flow rate of 300 nL/min. The acquisition parameters for the MS experiments are provided in the **Supplementary Data**.

### Data Processing and Database Searching

Thermo RAW files were processed using the latest version of Xcalibur (Thermo Fisher Scientific). Mass spectral peaks were automatically identified by the software using default settings and filtered to include only peaks with charge states between +2and +7 m/z. Spectral data were converted into .mgf files using MSconvert (ProteoWizard) or Mascot Distiller (Matrix Science, London, UK). The data were searched for peptide identification using Mascot (Matrix Science, London UK). Tandem MS data were searched against the latest version of a combined Human (taxonomy ID 9606) and Enterovirus (taxonomy ID 12059) databases downloaded from the latest UniProt database (https://www.uniprot.org/).

The following search parameters were used: precursor mass tolerance was set to 10 ppm and fragment mass tolerance was set as 0.08 Da. Enzyme was set as trypsin with two missed cleavages permitted. Carbamidomethylation of cysteine was set as a fixed modification and oxidation of methionine, deamidation of asparagine and glutamine, and protein N-terminal acetylation (protein N-Term) were set as variable modifications. The Mascot decoy database function was enabled, and the false discovery rate was set at < 1%, while individual ions scores >13 indicated identity or extensive homology (p<0.05). Only bold red peptides were considered in the protein identifications. A bold red match is the highest scoring match to a particular query listed under the highest scoring protein containing that match. Complementary analyses were performed using Protein Pilot (ABSciex), Pinnacle (Optys Tech Corporation) and Scaffold DIA (Proteome Software). Peptides that surpassed significant threshold levels in these analyses were included for protein identifications.

### Bioinformatics Analysis

BLAST annotation of identified viral proteins was performed using the BLASTP (https://blast.ncbi.nlm.nih.gov/). Sequence similarity search with an E-value threshold set at 1E-03 was carried out without taxonomical restriction against non-redundant protein sequences in the National Center for Biotechnology Information (NCBI) database. A search of the conserved domain (CD) of proteins with the Batch CD-Search tool of NCBI server was performed to support BLAST annotations.

### Immunoblot Detection of Virus VP1 Expression

Normalized (40 µg) of total protein lysates from the nPOD cases that were used for MS analysis were separated by SDS-PAGE and transferred to PDVF membranes and processed using standard Western blot methods. The membranes were incubated with the DAKO Clone 5D8/1 anti-VP1 mouse monoclonal antibody and anti-human GAPDH primary antibodies (sc-25778, Santa Cruz Biotechnologies) at 4°C overnight. The membranes were washed extensively with 0.1% Tween-20 in PBS before incubation with donkey anti mouse IRDye 800 and donkey anti goat IRDye 600-conjugated secondary antibodies. The membrane was washed extensively with 0.1% Tween-20 in PBS, before detection of VP1 and GAPDH as loading control using a LiCor Odyssey infrared Imager (LiCor, Lincoln, NE).

### Quantitation and Statistical Analysis

Quantitation methods and statistical methods for proteomic analysis are described in the relevant sections.

## Results

As part of a comprehensive assessment of viral proteins and RNA in pancreas tissue from donors with/without type 1 diabetes undertaken by the nPOD-Virus group, we examined the presence of viral proteins using qualitative and quantitative proteomics. The experimental strategy and workflow are summarized in **Figure 1**. These mass spectral data were also used to determine the presence of virus proteins/peptide, proteins that facilitate virus infection and proteins that are differentially expressed upon virus infection. Samples were analyzed from nPOD donors including those without diabetes, AAb+, with type 1 diabetes and with type 2 diabetes. For LC/MS/MS analysis, the samples were loaded and analyzed in randomized sequences.

**Figure 1.**
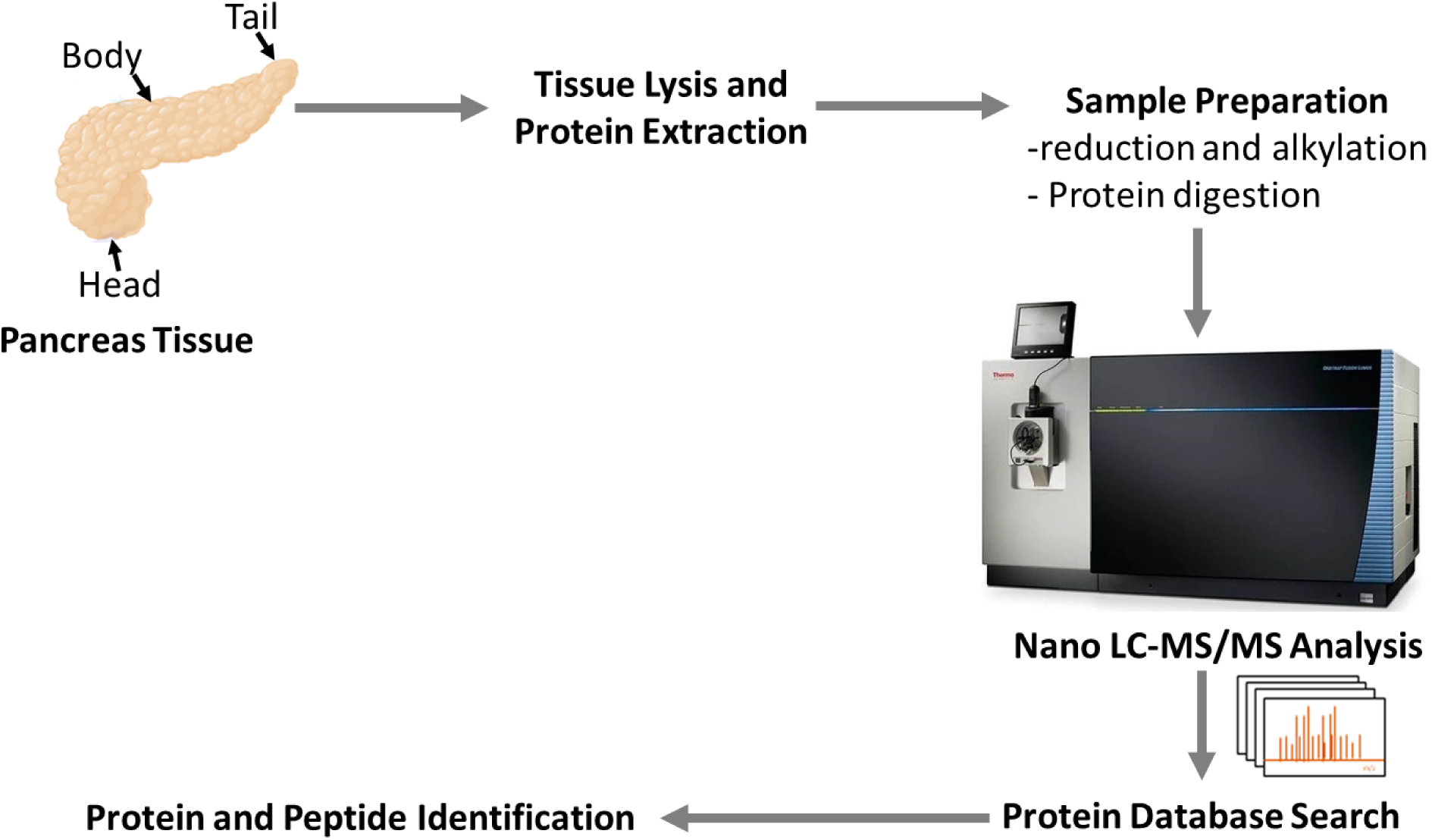
Experimental strategy. Workflow schematic for identification of viral proteins in nPOD pancreas tissues.

### Enterovirus Peptides are Identified in nPOD Pancreas Tissues

Enteroviruses, particularly CBVs, are environmental factors linked to type 1 diabetes by earlier studies [22–29]. LC–MS/MS based proteomics was used to identify virus peptides/proteins in pancreas tissue from nPOD donors. Our analyses generated >1,000 individual mass spectra data files. A viral peptide was considered unambiguously identified in Mascot database searches if it was bold red, with a p<0.05, and expectation value lower than 0.05. The expectation score is the number of matches with equal or better scores that are expected to occur by chance alone (Matrix Science, London UK). The Mascot expectation score is directly equivalent to the E-value in a Blast search result. The lower the expectation value, the more significant the score. For a score that is exactly on the default significance threshold (p<0.05), the expectation value is also 0.05. We identified enterovirus peptides from different potential serotypes in 29 donors, 11 and 7 of which, respectively had type 1 diabetes or were AAb+. Thus, 11/25 (44%) donors with type 1 diabetes and 7/17 (41%) AAb+ donors carried virus proteins in the pancreas. Viral proteins were also found in 11/33 (33%) ND donors and 0/7 donors with type 2 diabetes. While this study was not powered to detect statistical differences using proteomics, the results demonstrate the principle that viral proteins can be found in the pancreas of organ donors, including those with or at-risk for type 1 diabetes. Results are summarized in **Table 1**. Some of the identified peptides are from viruses or viral serotypes that were previously associated with type 1 diabetes. These serotypes include Coxsackie A virus (CAV), coxsackie B virus (CBV), and echovirus (ECV). These serotypes are represented in 26/29 cases.

**Table 1.** Summary of nPOD cases phenotypes and potential viral serotypes identified by LC-MS-MS. The serotype acronyms are: CVA-Coxsackievirus A, CVB-Coxsackievirus B, E – Echovirus, EV-Enterovirus, PEV-Parechovirus, PV-Polio virus, RV-Rhinovirus. The acronyms for disease phenotypes are: AAB+ - autoantibody positive, ND - Non-diabetes, T1D – type-1-diabetes. The serotypes that have been previously associated with type-1-diabetes are shown in bold. Some of the peptide sequences were not unique to a specific viral serotype and hence the inclusion of multiple potential serotypes under the fourth column.

| # | nPOD Case | Phenotype and other information | Potential Virus serotypes |
| --- | --- | --- | --- |
| 1 | 6084 | T1D, 4 years | **CVA, E9**, PV-3 |
| 2 | 6104 | ND | **CVA16, E11** |
| 3 | 6123* | AAB+ (GADA) | **CVA24, CVB1**, E9 |
| 4 | 6160 | ND | **CVA9, CVB1** |
| 5 | 6182 | ND | **E11**, RV-3 |
| 6 | 6195* | T1D, 5 years | **E9**, RV3 |
| 7 | 6198 | T1D, 3 years | **E6**, RV-3 |
| 8 | 6227 | ND | **EV-71**, RV3 |
| 9 | 6238 | ND | **EV71**, PV-1 |
| 10 | 6245 | T1D, 0.6 years | PV-1, PEV-9 |
| 11 | 6247 | T1D, 7 years | **CAV-16, EV-71**, RV-14, PV-3, PEV-9 |
| 12 | 6254* | ND | **CVB1, EV1, EV70** |
| 13 | 6263 | T1D, 21 years | **EV70**, PV-2 |
| 14 | 6267 | AAB+ (GADA, IA-2A) | PV-2, PV-3 |
| 15 | 6279 | ND | **E9, E30**, RV-1, RV-3 |
| 16 | 6289 | ND | **CVA-16**, RV-3, PV-3 |
| 17 | 6303 | AAB+ (GADA) | PV-1, PV-3 |
| 18 | 6318 | ND | **CVA-24**, PV-2 |
| 19 | 6325 | T1D, 6 years | **CVA-24, E9** |
| 20 | 6337 | T1D, 5 years | **CVB2, E9** |
| 21 | 6339* | ND | **CVA-16**, PV-1, PV-2 |
| 22 | 6362 | T1D diagnosed at death | **E5, E9** |
| 23 | 6366 | ND | **E9**, PV-1, PV-2 |
| 24 | 6367 | T1D, 2 years | **EV-70** |
| 25 | 6388 | AAB+ (GADA, mIAA) | **E9**, RV-2M, PEV-9 |
| 26 | 6397 | AAB+ (GADA) | **CAV16, CVB1, E9** |
| 27 | 6400 | AAB+ (GADA) | **CVB3, E6, E9**, RV-1, PV-3 |
| 28 | 6504 | T1D, 0.6 years | **E1, EV-71**, RV-3 |
| 29 | 6429 | AAB+ (GADA, mIAA) | **CAV21, CVB1** |

Figures 2A-B show representative MS/MS spectra from viral peptides and virus serotypes that were identified in some of the pancreas samples from nPOD donor 6160. The peptides are identified with well annotated b and y ion fragment series. The annotated ions in the spectrum and are shown in red in the corresponding fragmentation ion tables. Figure 2A shows a tandem mass spectrum identifying a VP1 peptide with amino acid sequence NVNFNPTGVTTTR. Figure 2B shows the spectrum for a VP4-VP2 peptide with sequence SMPALNSPSAEECGYSDR. It is notable that 10/29 donors with virus peptides shown by LC/MS/MS also tested positive for VP1 staining by immunohistochemistry or for enterovirus RNA by RT-PCR (see accompanying articles by Richardson et al. and Laiho et al). The detection of virus markers by these orthogonal methods provides complementary evidence for the presence of viruses in pancreata from nPOD donors; of note, full matching of results across methods cannot be expected due to the rarity of virus-positive cells and variability in sampling.

**Figure 2.**
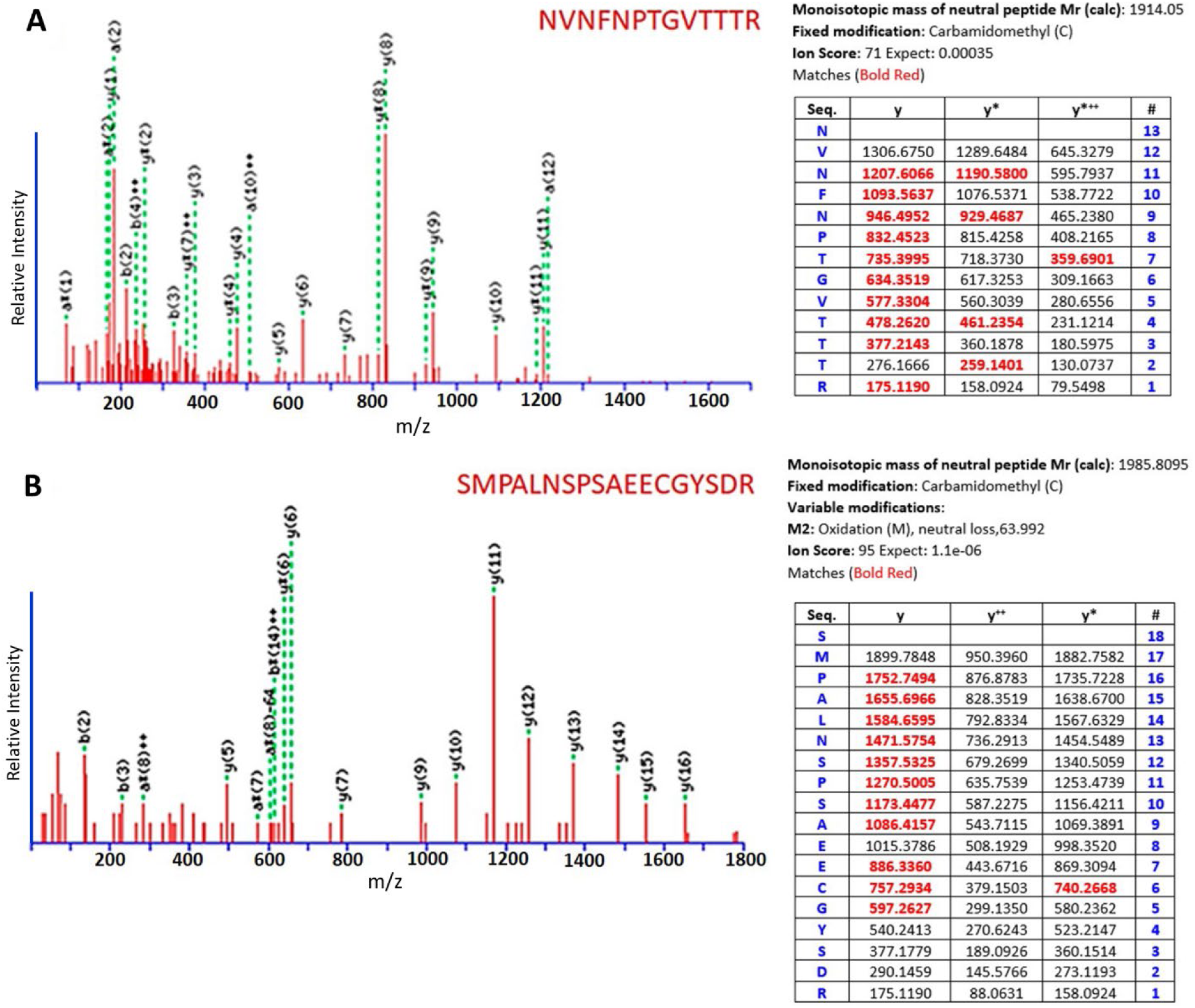
Representative MS/MS spectra and ion assignments for CVB1 peptides from nPOD case 6160 pancreas tissue. **A.** MS/MS fragmentation data of VP1 peptide with sequence NVNFNPTGVTTTR. **B.** MS/MS fragmentation data of VP4-VP2 peptide with sequence SMPALNSPSAEECGYSDR. In both panels A and B, the y-axis show the m/z of MS^2^ tandem mass spectra data with annotated primary b and y ion series of the fragments that are used to identify peptide sequences, where b ions represent fragments cleaved on the N-terminal end of the peptide bond and y ions representing fragments cleaved on the C-terminal end of the peptide. The annotated b and y ion series unambiguously identifying the two peptides. Each series contains contiguous y-ion fragments as well as Mascot ion scores and significant expect scores indicating a confident sequence match. Each figure shows the relative intensity of the individual fragments from the mass spectrum shown in centroid profile in red solid lines; the green dotted lines show the annotation of identified fragments. The table associated with each figure lists: the amino acid sequence of the identified peptide in the first column labeled “Seq”, and starting from the N-terminus asparagine residue in the first row to the C-terminus arginine residue in the last row; the mass of the peptide sequence fragment ions are shown in the y columns, which can be mapped by the number on the right column (“#). This case was also positive for VP1 by IHC.

### BLAST Analysis of Identified Virus Peptides

We performed Basic Local Alignment Search Tool (BLAST) analysis of the VP1 peptide with sequence NVNFNPTGVTTTR identified in nPOD donor 6160 to find regions of similarity between their primary amino acid sequences and those in the non-redundant protein sequences NCBI database. These analyses integrated a search of the peptides’ conserved domains and were performed without taxonomy restrictions. The top hit was polyprotein from Coxsackievirus B1 which was identified with 13/13 (100%) identities and 13/13 positives (100%) with an expectation score of 5e-04. A significant number of all the identified peptides in the database were highly homologous with a few exceptions where VP1 from Echovirus 12 was identified with a G>D substitution at position 8 of the peptide, NVNFNPTD*VTTTR with 12/13 (92%) identities and 12/13 positives (92%) with an expectation score of 0.012. Echovirus 5 was also identified with a sequence NVNFD*PTGVTTS*R that include two substitutions, a N>D at position 5 and a T>S at position 12. This corresponds to 11/13 (85%) identities and 12/13 positives (92%) with an expectation score of 0.048. **Figure 3A and 3B** shows alignments and the Tree view of the search produced using BLAST pairwise alignments and were analyzed using the Fast Minimum Evolution and Neighbor Joining methods. **Table 2**. Summarizes the Blast analysis results of the lineage of the peptide in the NCBI database.

**Figure 3.**
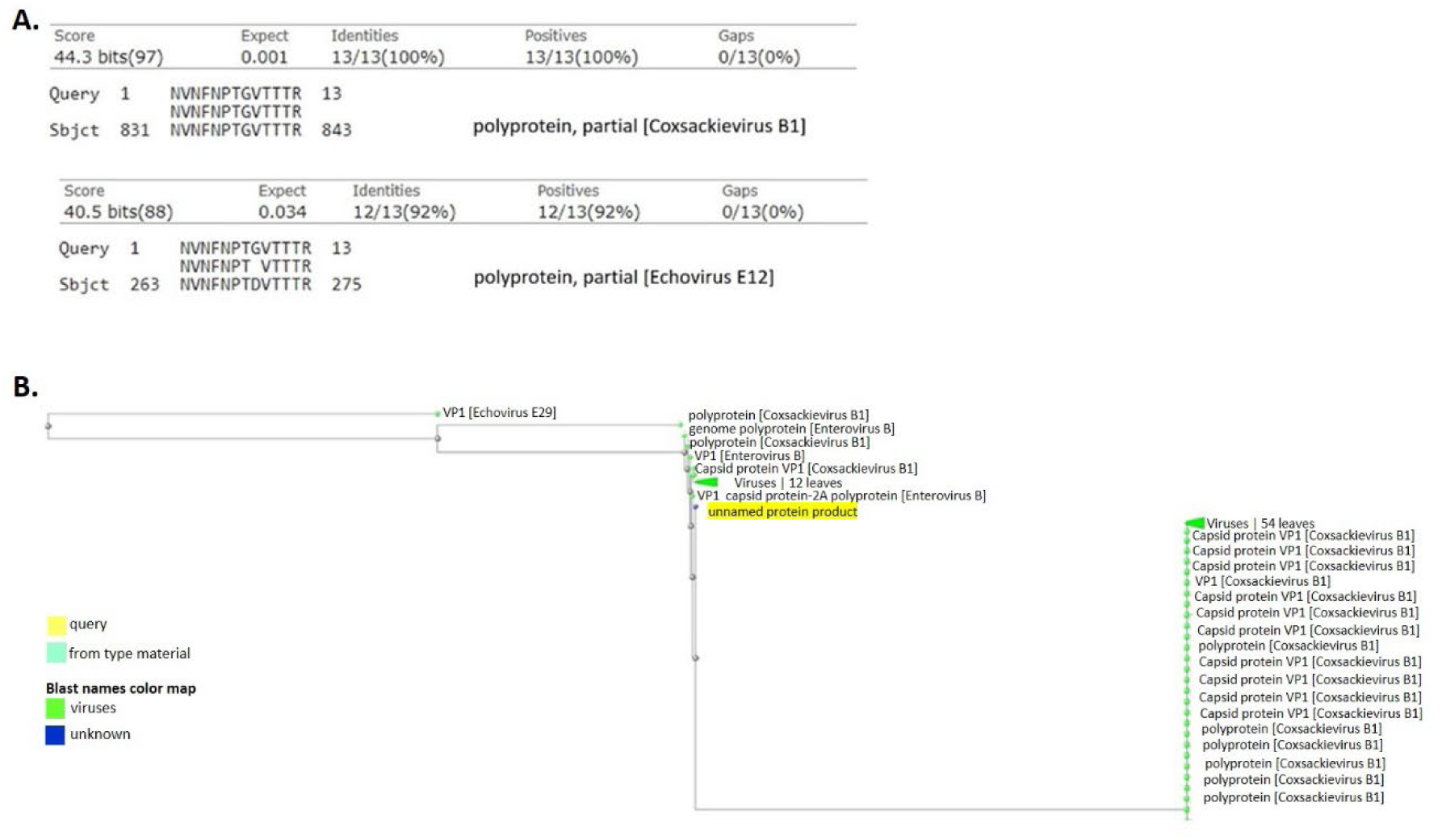
Blast analysis of VP1 peptide with sequence NVNFNPTGVTTTR from nPOD case 6160. **A.** Pairwise alignments showing the identification of VP1 from CVB1 with the highest probability score. Other significant alignments are for Echovirus 5 and Enterovirus B93. **B.** Tree view analysis of the alignments using the Neighbor Joining methods showing the lineage of the virus serotypes.

**Table 2.**
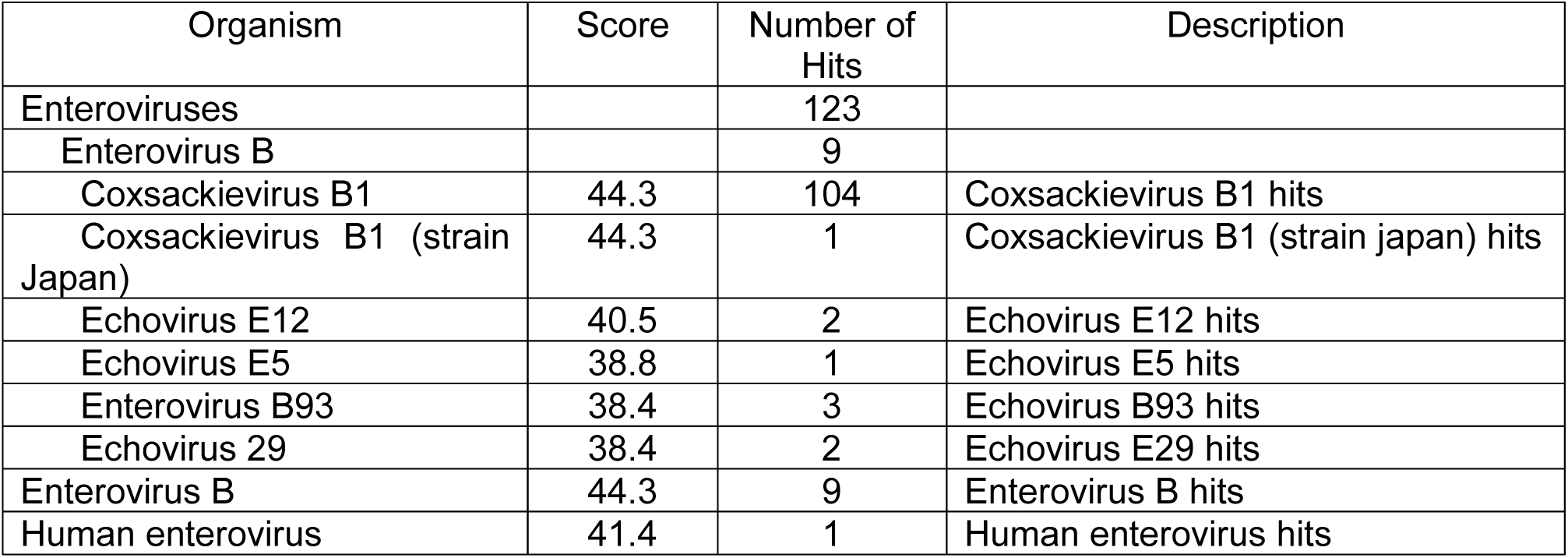
Summary of the taxonomy lineage report from the BLAST search results using the peptide NVNFNPTGVTTTR identified in nPOD case 6160; the number of organisms that are identified and the number of protein hits in the database are provided.

### Western blot Detection of VP1 in nPOD Pancreas Tissues

A sub-set of nPOD donors were selected for LC/MS/MS proteomic analyses based on positive enterovirus VP1 signals by IHC on FFPE tissue sections, and positive signals for HLA Class I hyper-expression and insulin using corresponding fresh frozen OCT embedded tissues sections. Corresponding control tissues were selected based on the absence of VP1 signals and low expression of HLA Class I molecules by islet cells. Based on these criteria, 30 µM pancreatic tissue slices from fresh frozen OCT embedded tissue were obtained from 8 ND donors, 3 AAb+ donors, and 7 donors with type 1 diabetes; tissues were used for protein isolation and LC/MS/MS and Western blot analysis. The demographic and clinical features of these cases are provided in **Supplementary Table 1**. Where available, 40 µg of total protein lysates from these donors were used for MS analysis, separated by SDS-PAGE, transferred to PDVF membranes, and processed using standard Western blot methods. The PVDF membranes were probed to detect virus VP1 protein and host GAPDH as protein loading control, using a LiCor Odyssey infrared Imager (LiCor, Lincoln, NE).

Figure 4 shows Western blot results. We did not obtain sufficient material for Immunoblot analysis for 3 donors (6401, 6367 and 6405). The amounts of GAPDH detected in the other 15 samples are similar. A VP1 signal is observable at variable amounts in some of the cases including 6254, 6339, 6401 and 6429. The specificity of the DAKO Clone 5D8/1 anti-VP1 mouse monoclonal antibody for enterovirus has been the subject of rigorous debate [30–32]. The detection of VP1 signals in a limited number of nPOD cases with VP-1 positivity by IHC supports that the antibody is specifically detecting VP1 and not host proteins, as has suggested previously. Non-specific detection of host proteins would have given uniformly detected signals across all the samples, similar to GAPDH albeit at presumably low intensities. The detection of enterovirus VP1 signals by immunoblot provides evidence for the presence of the virus in some of the nPOD cases. These results validate the LC/MS/MS using an independent orthogonal method.

**Figure 4.**
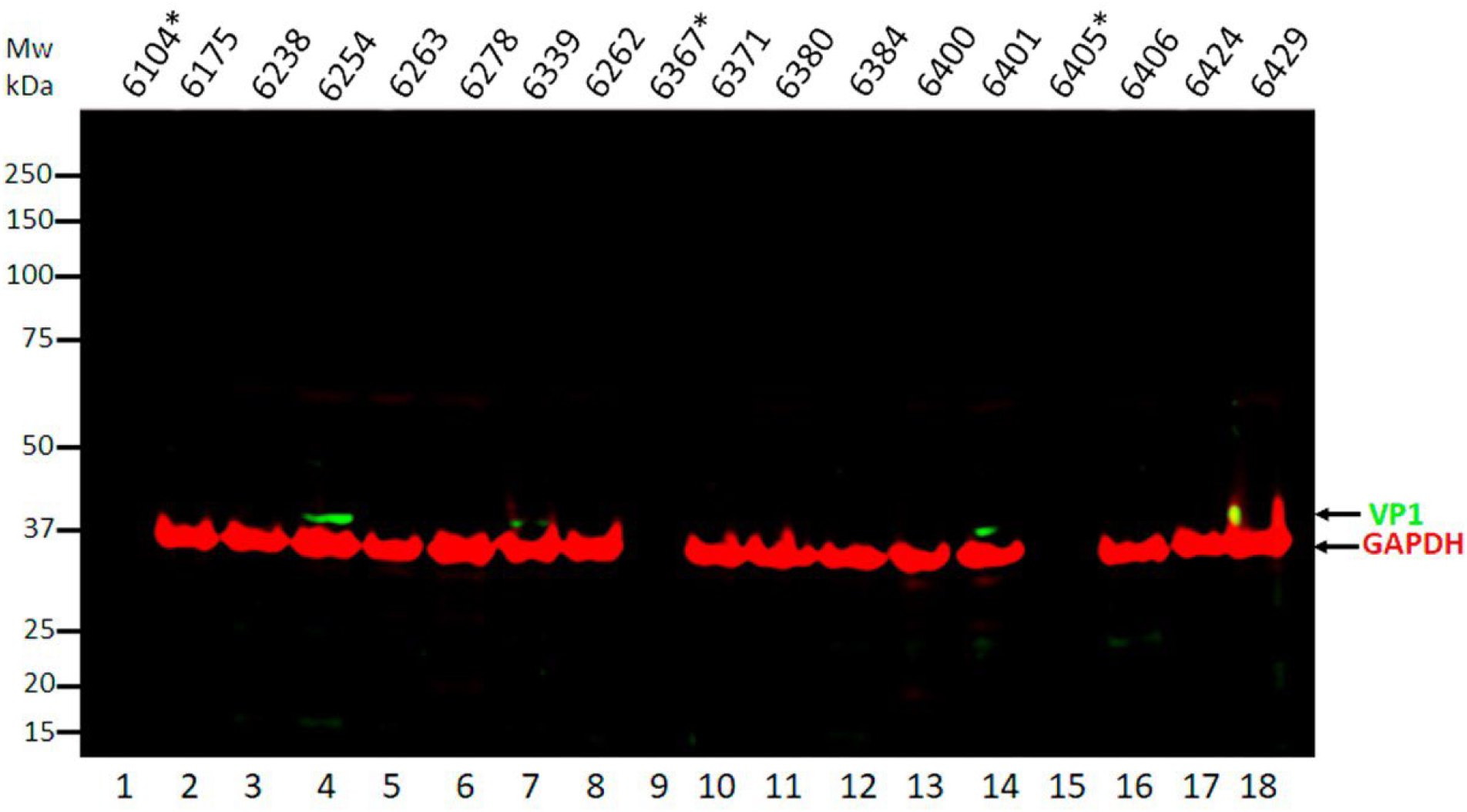
Normalized protein concentrations from 18 nPOD cases were used in immunoblots to evaluate the expression of enterovirus VP1 protein. GAPDH was used as a loading control for the experiments. Sufficient protein material was not available for three nPOD cases 6401, 6367 and 6405. Sample loading buffer was loaded in the three lanes that are marked with asterisks *.

Additional Western blot analysis using total lysates from 23 fresh frozen pancreas tissue chunks and OCT tissue samples were performed for four nPOD cases 6227 (ND), 6123 (AAb+), 6084 and 6195 (type 1 diabetes). In these analyses, clear VP1 bands are detectable in cases 6123 and 6195 in the OCT samples. LC/MS/MS data also identify virus peptides in these samples (**Figure 5**). **Figures 6a** and **6b** show tandem MS data identifying a VP1 peptide with sequence QAVEGAIGR from Echovirus 9 and a potential 2C peptide with sequence ITDSLEVLFQGPVYK from human rhinovirus 3 . These corresponding LC/MS/MS analyses data identify VP1 signals in total lysates from the fresh flash frozen cases, including 6160 and 6151 **(Figure 7)**.

**Figure 5.**
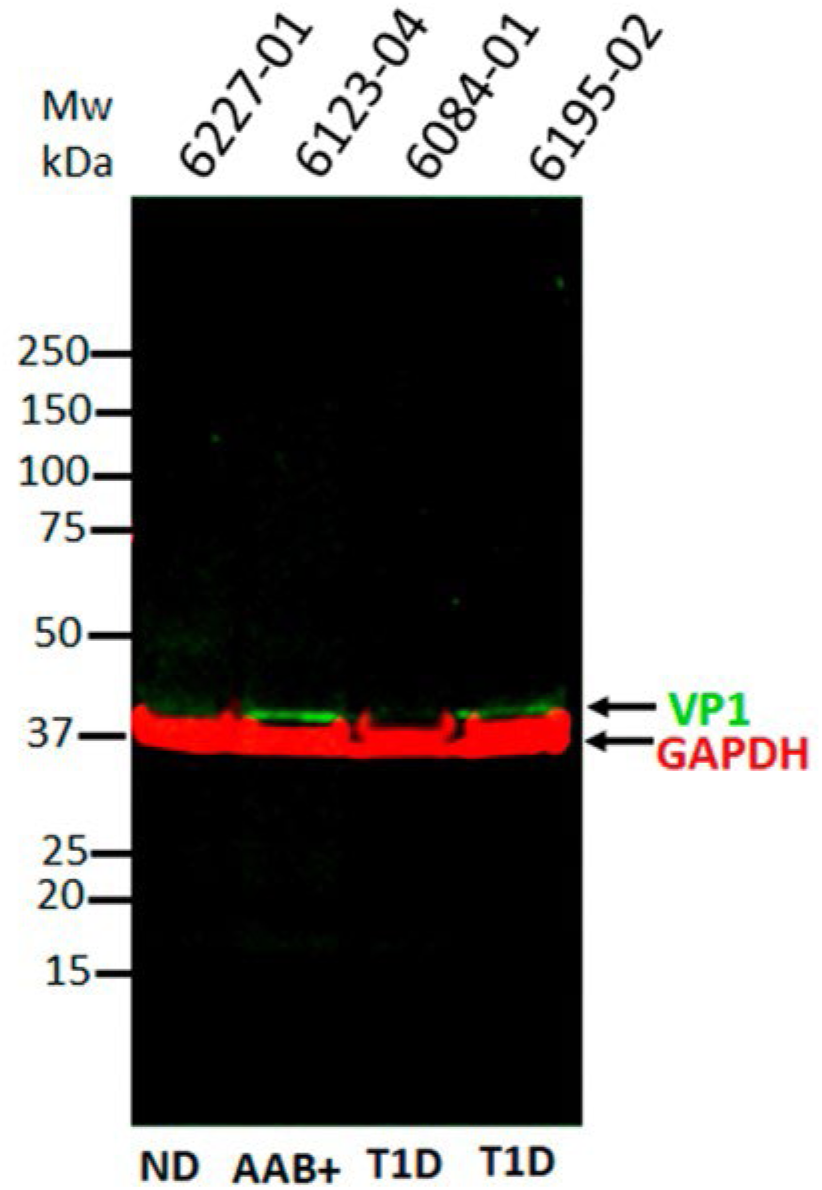
Normalized protein concentrations from four disease-stratified nPOD cases were used in immunoblots to evaluate the expression of VP1. GAPDH was used as a loading control for the experiments. ND denotes non-diabetes; AAB+ denotes autoantibody positive and T1D, type 1 diabetes.

**Figure 6.**
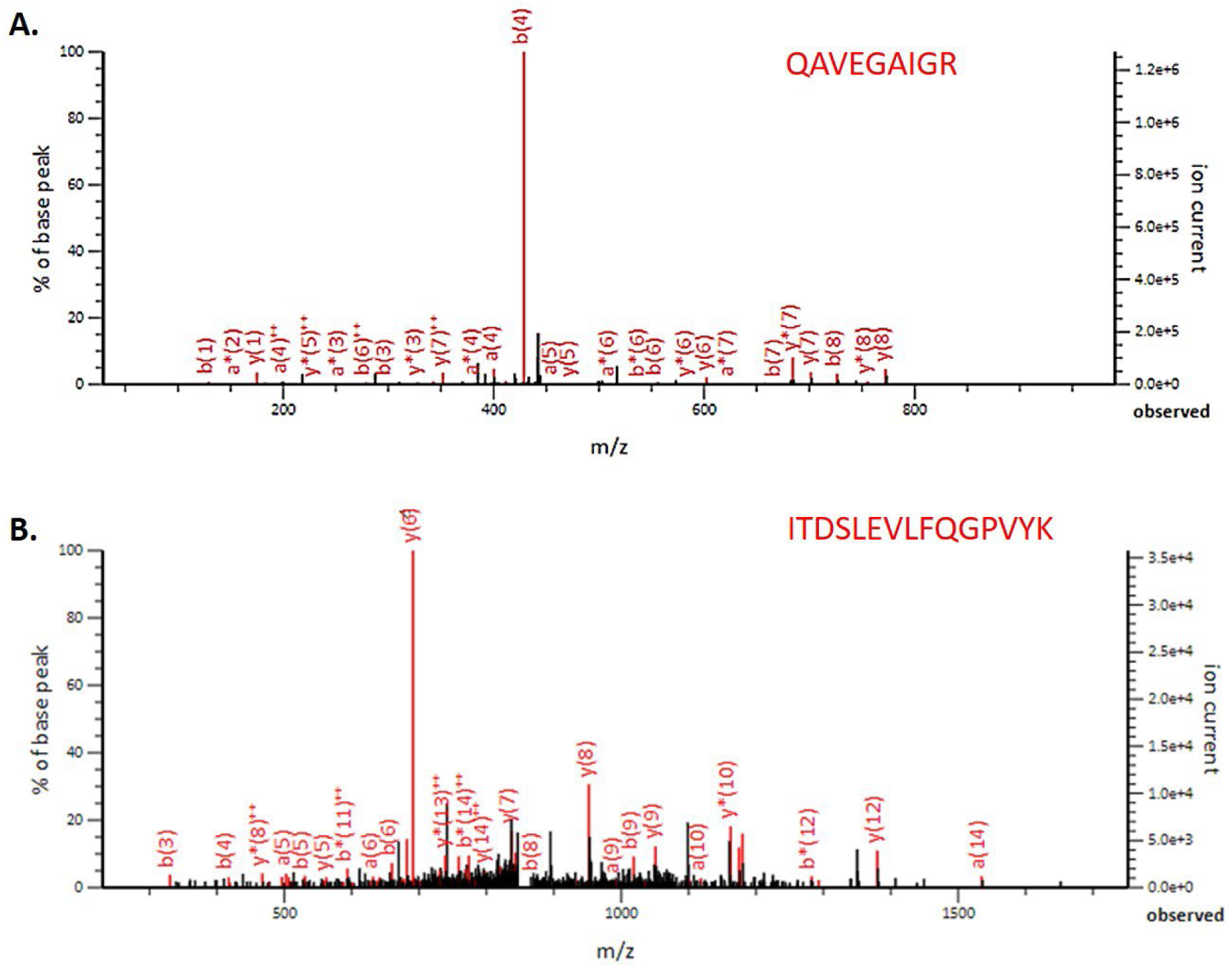
Tandem mass spectra of virus peptides identified from nPOD case 6195. **A.** MS/MS fragmentation data and identification of Echovirus 9 VP1 peptide with sequence QAVEGAIGR. **B**. MS/MS fragmentation data and identification of 2C peptide with sequence ITDSLEVLFQGPVYK from human rhinovirus 3. The y-axis show the m/z of MS^2^ tandem mass spectra data with annotated primary b and y ion series of the fragments that are used to identify peptide sequences, where b ions represent fragments cleaved on the N-terminal end of the peptide bond and y ions representing fragments cleaved on the C-terminal end of the peptide.

**Figure 7.**
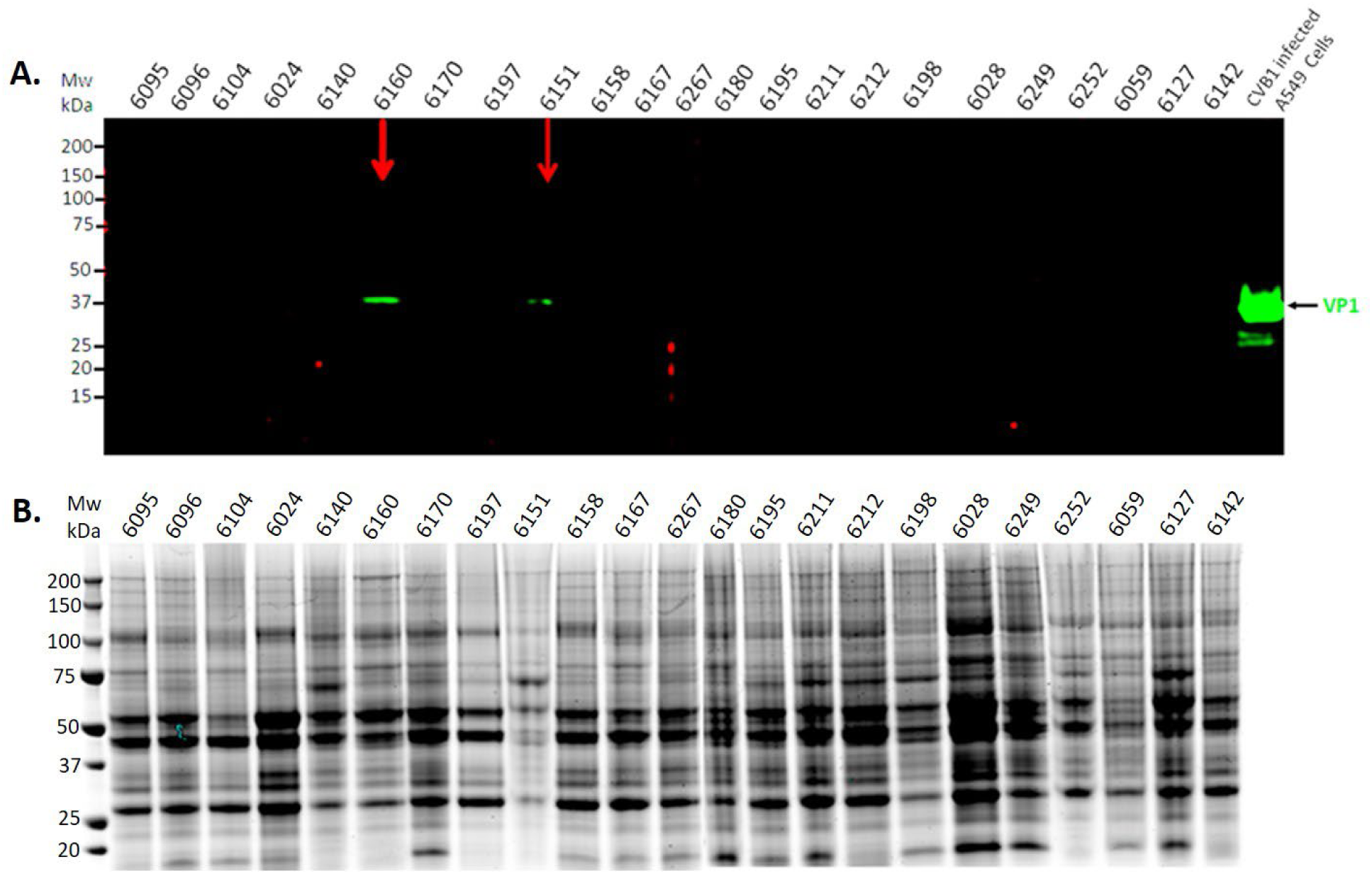
SDS-PAGE and Western Blot Analysis of Pancreas Tissue Lysates. **A.** Normalized protein concentrations from 23 nPOD cases were used in immunoblots to evaluate the expression of enterovirus VP1. CVB1 infected A549 cells were used as positive control for VP1 detection. Cases 6160 and 6151 that show significant detectable levels of VP1 are marked with red arrows. **B.** Colloidal Coomassie stained gel for the 23 nPOD cases to show complexity of fractionated proteins.

### Comprehensive analysis of nPOD case 6160 by Gel-LC-MS-MS

In Western blot analysis, VP1 bands at the expected molecular weight (37 KDa) is detected for donors 6160 and 6151, which are comparable to the VP1 signal in the control A459 cell lysates **(Figure 7)**. nPOD case 6160 demonstrates a relatively higher abundance detection of enterovirus VP1 compared to all the other samples (Figure 7). Since there is a very limited number of specific antibodies available for the detection of other virus proteins, we used Gel-LC/MS/MS for comprehensive proteomic analysis. Fresh frozen pancreas tissues were lysed by homogenization and sonication in RIPA buffer. Protein concentrations were determined by BCA before fractionation by SDS-PAGE. After gel separation and staining/destaining, gel slices were excised, and subjected to in-gel reduction, alkylation, and trypsin digestion. Tryptic peptides were processed and subjected to LC-MS/MS analysis. Twenty unique Coxsackievirus B1 peptides were identified, including peptides from VP1, VP2 and VP4 structural proteins and the 2C non-structural protein that induces and associates with structural rearrangements of intracellular membranes in addition to exhibiting RNA-binding, nucleotide binding and NTPase activities. The identified peptides are shown in **Table 3**. **Figures 2A-B** shows tandem MS spectrum for VP1 and VP4-VP2 peptides with sequences NVNFNPTGVTTTR and SMPALNSPSAEECGYSDR, respectively. Additional Mascot database data identifying the VP1 peptide with sequence SESSIENFLCR and peptide 2C with sequence SVATNLIGR from this donor are shown in **Figures 8A** and **B**.

**Figure 8.**
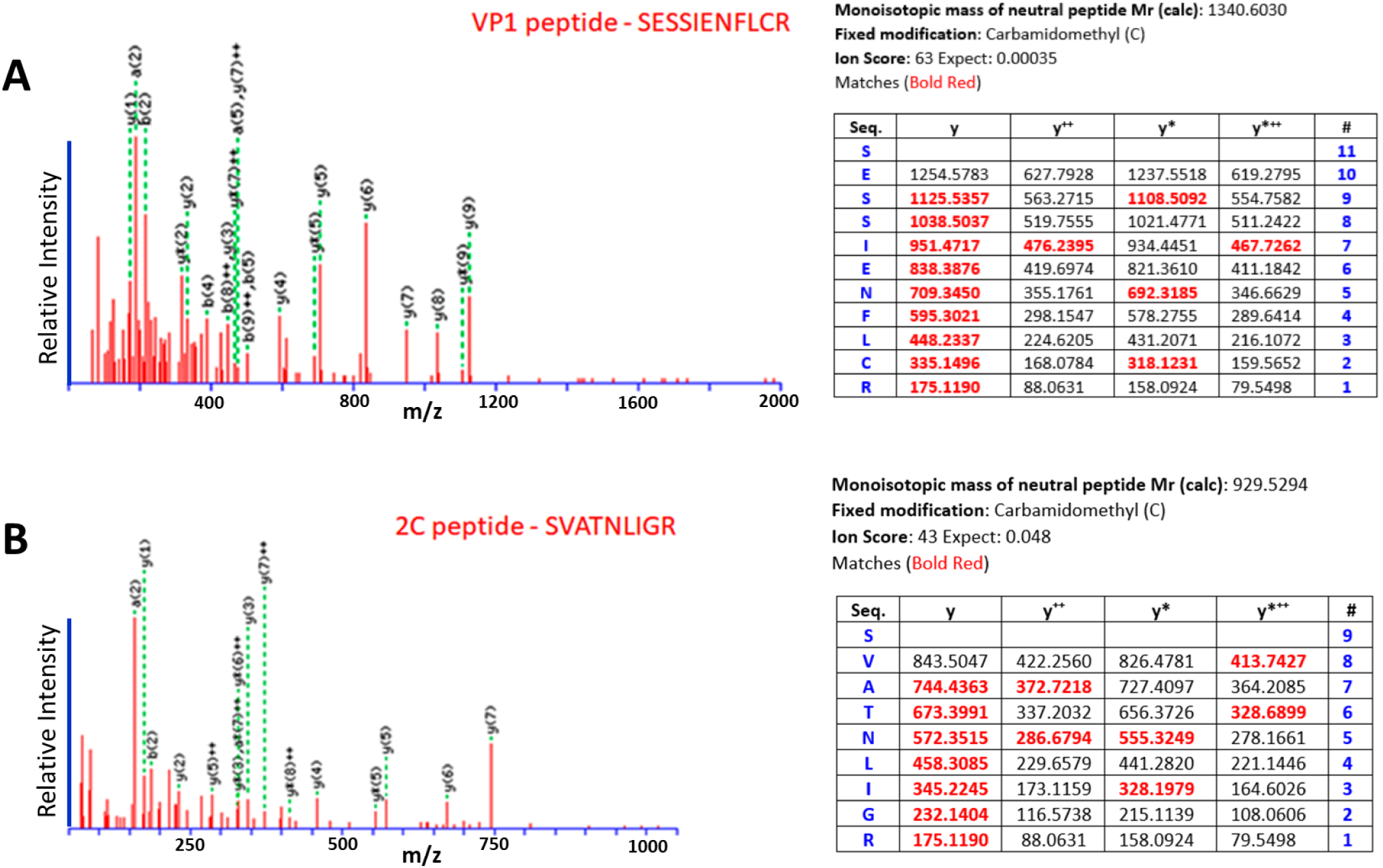
Tandem mass spectra of virus peptides identified from nPOD case 6160. **A.** MS/MS fragmentation data and identification of Coxsackievirus B1 VP1 peptide with sequence SESSIENFLCR. **B.** MS/MS fragmentation data and identification of 2C peptide with sequence SVATNLIGR from CVB1. In both panels A and B, the y-axis show the m/z of MS^2^ tandem mass spectra data with annotated primary b and y ion series of the fragments that are used to identify peptide sequences, where b ions represent fragments cleaved on the N-terminal end of the peptide bond and y ions representing fragments cleaved on the C-terminal end of the peptide. . Each figure shows the relative intensity of the individual fragments from the mass spectrum shown in centroid profile in red solid lines; the green dotted lines show the annotation of identified fragments. The table associated with each figure lists: the amino acid sequence of the identified peptide in the first column labeled “Seq”, and starting from the N-terminus asparagine residue in the first row to the C-terminus arginine residue in the last row; the mass of the peptide sequence fragment ions are shown in the y columns, which can be mapped by the number on the right column (“#).

**Table 3.**
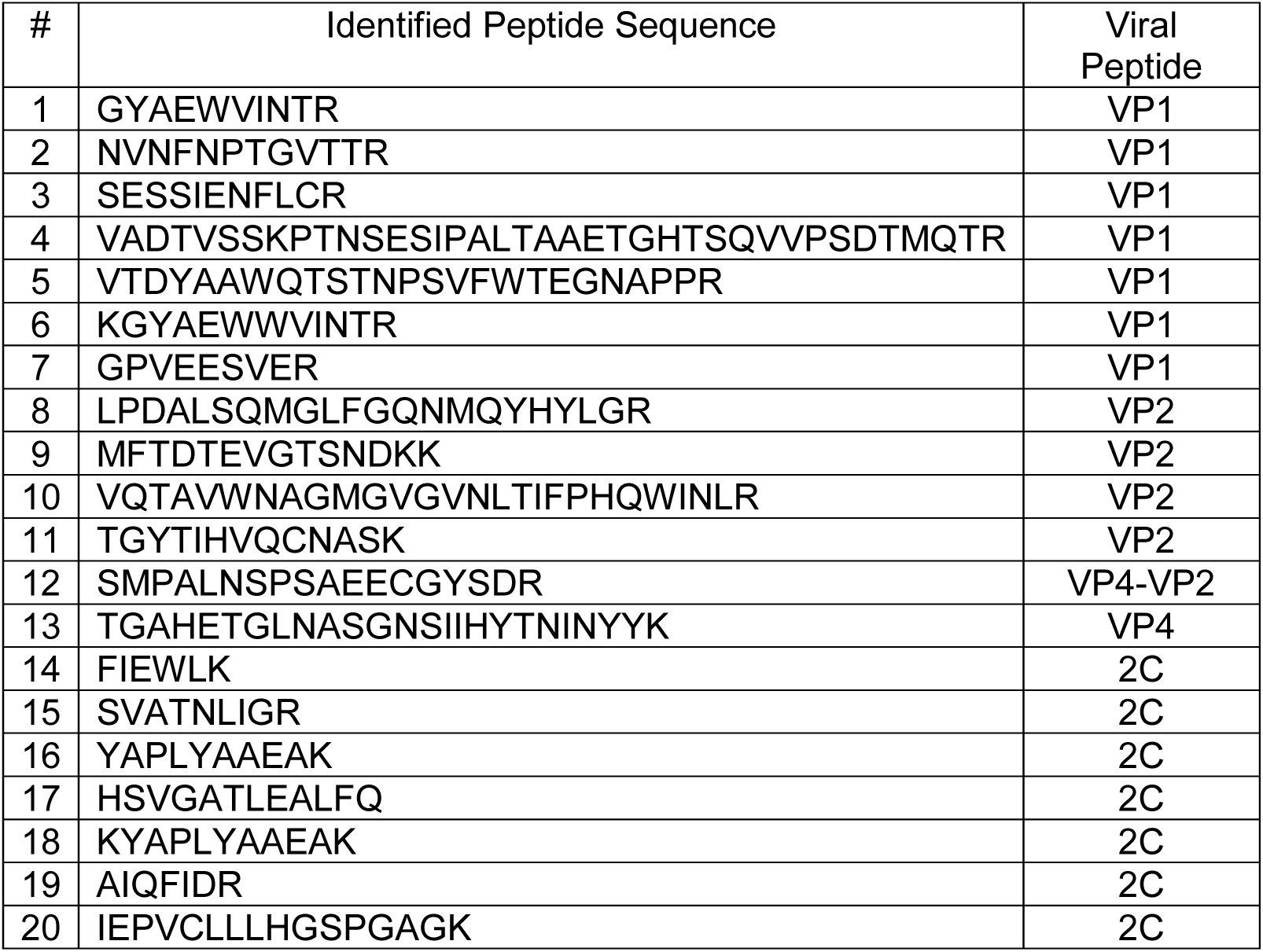
Sequences of Coxsackievirus B1 peptides identified by Gel-LC-MS-MS in nPOD case 6160.

### Virus Infection Mediating Proteins are Identified in nPOD Pancreas

Enteroviruses use various receptor molecules to invade their host cells. The main receptor for CBVs is the tight junction protein Coxsackie–adenovirus receptor (CAR), which belongs to the large family of adhesion molecules [33–35]. An objective of this study was to use LC/MS/MS to identify CAR protein in nPOD pancreas tissues. We identified CAR protein in nPOD pancreata using both non-targeted and targeted LC/MS/MS approaches. Figure 9 shows MS/MS spectra for a CAR peptide with sequence TQYNQVPSEDFER, amino acid positions 316-328 in the primary sequence of the protein. This representative tandem MS spectra is from nPOD case 6195, a patient with type 1 diabetes with a disease duration of 5 years. The peptide is identified unambiguously with well annotated b and y ion fragment series. These ions are shown and depicted in red in the adjacent fragmentation table. Previous studies have demonstrated that the frequency of CAR expression is higher in pancreas of T1D tissues and autoantibody-positive non-diabetic donors compared with controls [36].

**Figure 9.**
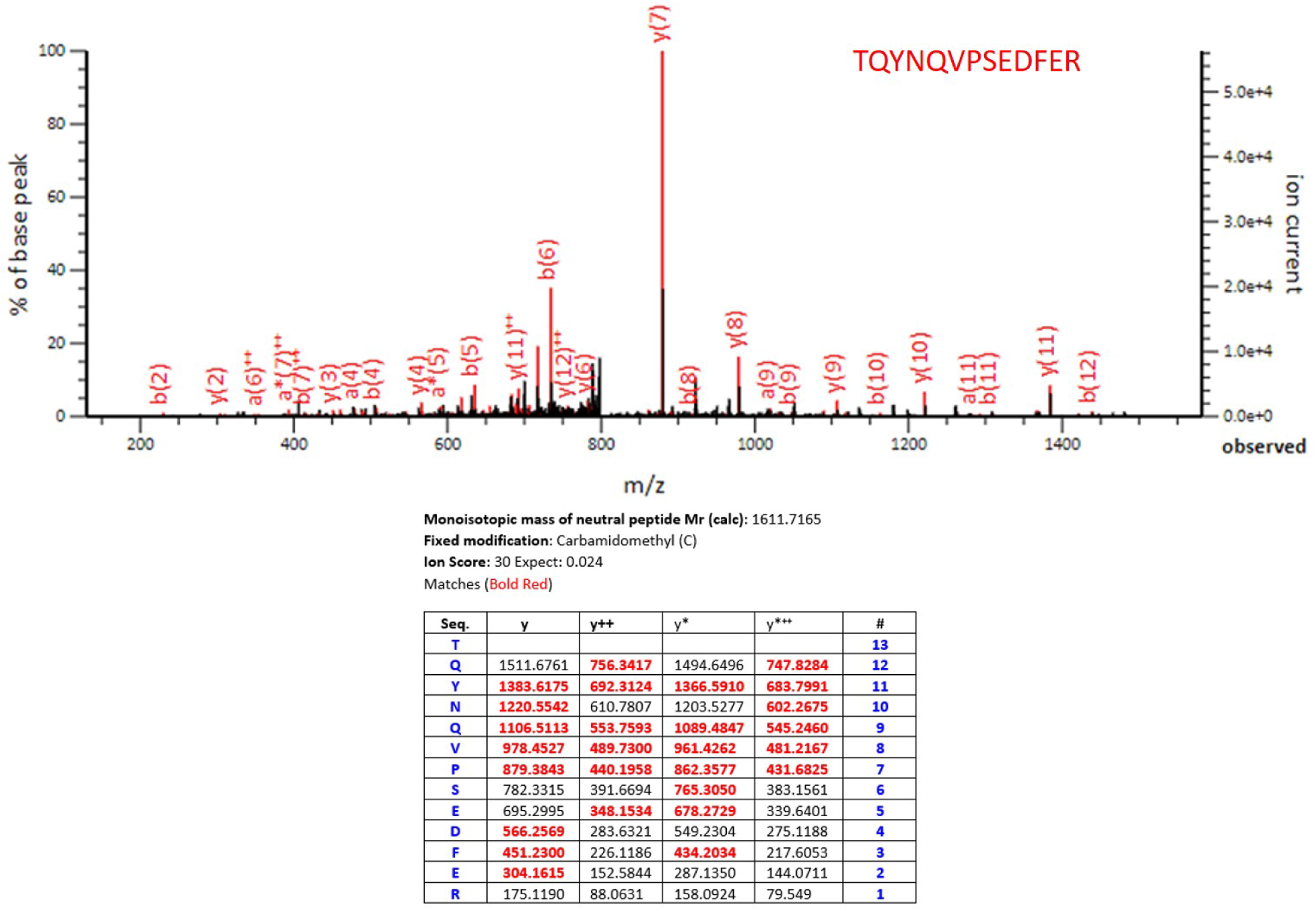
Tandem mass spectrum of Coxsackievirus and adenovirus receptor peptide TQYNQVPSEDFER from nPOD case 6195 unambiguously identifying the protein. The table associated with each figure lists: the amino acid sequence of the identified peptide in the first column labeled “Seq”, and starting from the N-terminus asparagine residue in the first row to the C-terminus arginine residue in the last row; the mass of the peptide sequence fragment ions are shown in the y columns, which can be mapped by the number on the right column (“#).

## Discussion

Enteroviruses have been previously linked to type 1 diabetes as environmental triggers and risk factors for islet autoimmunity [37–43]. In the context of the coordinated investigation conducted by the nPOD-Virus group, our task was the identification of viral peptides in organ donor pancreata using mass spectrometry-based proteomics. Our goal instead was to apply advanced proteomics to enable the detection of enterovirus proteins, to provide additional evidence of their presence in the pancreas, and inform about their nature, to complement the approaches implemented by the nPOD-Virus Group. Our approach is not practical as screening tool to demonstrate an association with disease or a specific donor group, but rather it allows identification of specific virus and virus serotypes directly in pancreas, as such evidence is limited. Moreover, the proteomic approach has not been used for this purpose previously. We have shown that mass spectrometry-based proteomics is a sensitive technique for the identification of pathogens in addition to other techniques such as microscopy-based antigen detection and nucleic acid-based approaches, such as PCR/RT-PCR [44]. Here, we used multiple LC/MS/MS strategies to identify enterovirus peptides in pancreas tissues. Our study provides evidence for the presence of enterovirus proteins in these tissues and allows for identification of viruses and virus serotypes. It is important to note that the viral proteins are present at very low stoichiometric levels compared to the host proteins. This is evidenced by the low overall intensities of the virus peptide ions at the MS1 and MS2 levels. Our data provides evidence that human pancreata may be infected be enteroviruses as they harbor various enterovirus proteins.

The detection of enterovirus proteins not exclusively among donors with type 1 diabetes (44%) but also among donors without diabetes (33%) is consistent with the high prevalence of these infections in the general population; however, we did not detect enterovirus proteins among 7 donors with type 2 diabetes. The number of donors that is possible to examine by proteomics is limited and preclude sufficiently powered case-control studies seeking to demonstrate an association of enterovirus infections with disease. We note that this study is part of a broader effort to investigate enterovirus infections in the pancreas, in relation to type 1 diabetes, by the nPOD-Virus Group. Collectively, the group has used a multitude of approaches (immunohistochemistry, proteomics, RNA sequencing and RT-PCR, in situ hybridization). A joint analysis of the data cumulatively generated by the nPOD-Virus group is presented in another manuscript in which our data were integrated in a more comprehensive analysis of association.

Despite the above limitations, our proteomic analysis provides independent evidence of enterovirus infections in the human pancreas, and the detection of non-structural proteins which are only synthesized during viral replication supports the concept that viruses had replication potential. Enteroviruses are known to disappear several months after the onset of diabetes, especially in fulminant type 1 diabetes and typical acute-onset T1D. However, they persist in cases of slowly progressive T1D. The low levels of enterovirus proteins detected in our study may be consistent with low-grade replication and persisting infection. Among the 25 donors with type 1 diabetes we examined, 13 had evidence of enterovirus proteins in their pancreas. There was no difference among enterovirus positive and negative donors with type 1 diabetes when comparing the age at passing, age of onset, and disease duration, as shown in **Supplemental Table 3**. Overall, the novel application of MS methods to identify virus and enterovirus serotypes from human pancreas is important to understand the biology underlying viral infections and their potential role in the development of type 1 diabetes. This study provides strong independent data on the potential role of enteroviruses in type 1 diabetes in support of other nPOD-V Group studies [45–48], the Diabetes Virus Detection (DiViD) Group studies [46–51]. In addition, this study also provides evidence for published data on the role of enterovirus on persistent infections in T1D [52–54], and the persistence of Coxsackie virus infections in pancreas upon 5’ terminal deletion [55]. (Tracy et al. J. Med. Virol. 2015; 87(2):240-247).

Overall, the novel application of MS methods to identify virus and enterovirus serotypes from human pancreas is important to understand the biology underlying viral infections and their potential role in the development of type 1 diabetes. The identification of specific molecules may provide targets for therapeutic interventions and some of the molecules may be potential prognostic markers for future development of type 1 diabetes.

## Abbreviations

Aab: autoantibody
CAR: coxsackie-adenovirus receptor
CVB: coxsackievirus B
LC: liquid chromatography
LCM: laser capture microdissection
MS: mass spectrometry
ND: No diabetes
nPOD: network for Pancreatic Organ donors with Diabetes
VP1: viral capsid protein 1

## Acknowledgements

We thank the families of the organ donors for the gift of pancreatic tissues.

## Funding

This research was performed with the support of the Network for Pancreatic Organ donors with Diabetes (nPOD; RRID:SCR_014641), a collaborative type 1 diabetes research project supported by grants from JDRF/ The Leona M. & Harry B. Helmsley Charitable Trust (3-SRA-2023-1417-S-B) and the Helmsley Charitable Trust (2018PG-T1D053, G-2108-04793). The content and views expressed are the responsibility of the authors and do not necessarily reflect the official view of nPOD. Organ Procurement Organizations (OPO) partnering with nPOD to provide research resources are listed at https://npod.org/for-partners/npod-partners/. The nPOD-Virus group was supported by JDRF grants (3-SRA-25-2012-516 and 3-SRA-2017-492-A-N awarded to A.P.).

## Data availability

The datasets analyzed during the current study are available from the corresponding author on reasonable request.

## Authors’ relationships and activities

The authors declare that there are no relationships or activities that might bias, or be perceived to bias, their work.

## Contribution of authors

**TCB**, acquisition, analysis and interpretation of data for the work. Final approval of the version to be published. **NLH**, acquisition, analysis and interpretation of data for the work. Final approval of the version to be published. **MAM**, design of the work, analysis and interpretation of data for the work, reviewing it critically for important intellectual content, final approval of the version to be published. **AP**, design of the work, reviewing it critically for important intellectual content, final approval of the version to be published. **JLN**, design of the work, analysis and interpretation of data for the work, reviewing the article critically for important intellectual content; and final approval of the version to be published. **JON**, design of the work, acquisition, analysis or interpretation of data for the work, drafting the article, reviewing it critically for important intellectual content and final approval of the version to be published.

## Supplementary Methods

### Mass Spectrometry Data Acquisition Methods

Acquisition of MS data was on the Q-Exactive MS were performed under the following: Full scans (m/z 350–1,700) at a resolution of 70,000 using an automatic gain control (AGC) target value of 3e6 and a maximum ion injection time (IT) of 100 ms. Tandem mass spectra were generated for up to 20 precursors by higher-energy collisional dissociation (HCD) using a normalized collision energy (NCE) of 28%. The dynamic exclusion was set to 35 s. Fragment ions were detected at a resolution of 17,500 with a 1e5u AGC and maximum IT of 50 ms.

The following acquisition parameters were used on the Orbitrap Fusion Lumos MS using data dependent acquisition (DDA) and data independent acquisition (DIA) methods. DDA acquisitions were under the following conditions: Full MS scans were performed at a resolution of 120,000, a maximum IT of 50 ms, AGC target value of 5e5, followed by MS2 events with a duty cycle of 2 s for the most intense precursors and with a 60 s with dynamic exclusion time. HCD scans were acquired with 35% NCE, with 1e5 AGC target, 0.25 activation Q, and 20 ms maximum injection time with a 1.3 m/z isolation width to inject ions for all available parallelizable time enabled. The MS2 scans were acquired in the Orbitrap under the following conditions; 1e5 AGC target, 0.25 activation Q and 100 ms maximum IT.

DIA analysis was performed on an Orbitrap Fusion Lumos MS under the following conditions: Mass acquisition range was from 400 to 900 m/z using 6 m/z quadrupole isolation windows, multiplexing these windows together randomly at the rate of 4 per scan. Maximum IT of 100 ms were used for both MS and MS/MS scans. AGC values were set to 4×10e5 for MS and 10e6 for MS/MS. HCD fragmentation was performed using 30% NCE with acquisition range between 200– 2000 m/z.

---

**Supplementary Table 1.** Donor phenotype and pancreas tissue samples analyzed by LC/MS/MS. The acronyms for disease phenotypes are: AAB+ - autoantibody positive, ND - Non-diabetes, T1D – type-1-diabetes.

| Study Number | Donor Type | Age Years | Sex | BMI (kg/m2) | C-peptide (ng/ml) | Duration | AAb | Race |
| --- | --- | --- | --- | --- | --- | --- | --- | --- |
| 6024 | ND | 21-25 | Male | 27.8 | 3.52 | NA | NA | Caucasian |
| 6028 | T2D | 31-35 | Male | 30.2 | 22.4 | 17 |  | Caucasian |
| 6046 | T1D | 16-20 | Female | 25.2 | nd | 8 | IA2A+<br>ZnT8A+ | Caucasian |
| 6052 | T1D | 11-15 | Male | 20.3 | 0.18 | 1 | IA-2A+,<br>mIAA+ | African American |
| 6059 | T2D | 16-20 | Female | 39.3 | 10.68 | 0.25 | NA | Hispanic/Latino |
| 6070 | T1D | 21-25 | Female | 21.6 | nd | 7 | IA-2A+,<br>mIAA+ | Caucasian |
| 6073 | ND | 16-20 | Male | 36 | 0.69 | NA | NA | Caucasian |
| 6080 | AAb++ | 56-70 | Female | 21.3 | 1.84 | NA | mIAA+<br>GADA+ | Caucasian |
| 6084 | T1D | 11-15 | Male | 26.3 | nd | 4 | mIAA+ | Caucasian |
| 6088 | T1D | 31-35 | Male | 27 | nd | 5 | GADA+<br>IA2A+<br>mIAA+<br>ZnT8A+ | Caucasian |
| 6090 | AAb+ | 0-5 | Male | 18.8 | 5.34 | NA | GADA+ | Hispanic/Latino |
| 6095 | ND | 36-40 | Male | 35.5 |  | NA | NA | Hispanic/Latino |
| 6096 | ND | 16-20 | Female | 18.8 | 2.97 | NA | NA | African American |
| 6099 | ND | 11-15 | Male | 30 | 5.37 | NA | NA | Caucasian |
| 6102 | ND | 41-45 | Female | 35.1 | 0.55 | NA | NA | Caucasian |
| 6103 | ND | 0-5 | Male | 16.8 | 0.98 | NA | NA | Caucasian |
| 6104 | ND | 41-45 | Male | 20.5 | 20.55 | NA | NA | Caucasian |
| 6113 | T1D | 11-15 | Female | 24.75 | nd | 1.58 | mIAA+ | Caucasian |
| 6123 | AAb+ | 21-25 | Female | 17.6 | 2.01 | NA | GADA+ | Caucasian |
| 6127 | T2D | 41-45 | Female | 30.4 | <0.08 | 10 | mIAA+ | Caucasian |
| 6128 | T1D | 31-35 | Female | 22.2 | nd | 31.5 | mIAA+ | Caucasian |
| 6137 | ND | 6-10 | Female | 24.2 | 12.13 | NA | NA | Hispanic/Latino |
| 6140 | ND | 31-35 | Male | 21.7 | 11.1 | NA | NA | Caucasian |
| 6142 | T2D | 26-30 | Female | 34.4 | 0.19 | 14 | mIAA+ | Caucasian |
| 6143 | T1D | 31-35 | Female | 26.1 | nd | 7 | IA2A+<br>mIAA+ | Caucasian |
| 6151 | AAb+ | 26-30 | Male | 24.2 | 5.49 | NA | GADA+ | Caucasian |
| 6158 | AAb++ | 36-40 | Male | 29.7 | 0.51 | NA | mIAA+<br>GADA+ | Caucasian |
| 6160 | ND | 21-25 | Male | 23.9 | 0.4 | NA | NA | Caucasian |
| 6167 | AAb++ | 36-40 | Male | 26.3 | 5.43 | NA | IA2A+<br>ZnT8A+ | Caucasian |
| 6170 | Aab+ | 31-35 | Female | 26.3 | 5.43 | NA | GADA+ | African American |
| 6175 | T1D* | 41-45 | Male | 19.9 | 3.48 | 12 | GADA+ | Caucasian |
| 6178 | ND | 21-25 | Female | 27.5 | 4.55 | NA | NA | Caucasian |
| 6180 | T1D | 26-30 | Male | 25.9 | nd | 11 | GADA+<br>IA2A+<br>mIAA+ | Caucasian |
| 6182 | ND | 0-5 | Male | 26 | 2.28 | NA | NA | Caucasian |
| 6184 | AAb+ | 46-50 | Female | 27 | 3.42 | NA | GADA+ | Hispanic/Latino |
| 6195 | T1D | 16-20 | Male | 23.7 | nd | 5 | GADA+<br>IA2A+<br>mIAA+<br>ZnT8A+ | Caucasian |
| 6197 | AAb++ | 21-25 | Male | 28.2 | 17.48 | NA | GADA+<br>IA2A+ | African American |
| 6198 | T1D | 21-25 | Female | 23.1 | nd | 3 | GADA+<br>IA2A+<br>mIAA+<br>ZnT8A+ | Hispanic/Latino |
| 6209 | T1D | 0-5 | Female | 15.9 | 0.1 | 0.25 | IA2A+<br>mIAA+<br>ZnT8A+ | Caucasian |
| 6211 | T1D | 21-25 | Female | 24.4 | nd | 4 | GADA+<br>IA2A+<br>mIAA+<br>ZnT8A+ | African American |
| 6212 | T1D | 16-20 | Male | 29.1 | nd | 5 | mIAA+ | Caucasian |
| 6213 | ND* | 21-25 | Female | 39.4 | 11.54 |  | NA | Caucasian |
| 6227 | ND | 16-20 | Female | 26.4 | 2.75 | NA | NA | Caucasian |
| 6228 | T1D | 11-15 | Male | 17.4 | 0.1 | 0 | GADA+<br>IA2A+<br>ZnT8A+ | Caucasian |
| 6234 | ND | 16-20 | Female | 25.6 | 6.89 | NA | NA | Caucasian |
| 6238 | ND | 16-20 | Male | 21.7 | 1.17 | NA | NA | African American |
| 6245 | T1D | 21-25 | Male | 23.2 | nd | 7 | GADA+<br>IA2A+ | Caucasian |
| 6247 | T1D | 21-25 | Male | 24.3 | 0.47 | 0.6 | mIAA+ | Caucasian |
| 6249 | T2D | 41-45 | Female | 32.2 | 4.17 | 15 | mIAA+ | Asian |
| 6252 | T2D | 16-20 | Male | 37.2 | 0.14 | NA | NA | Caucasian |
| 6254 | ND | 36-40 | Male | 30.5 | 6.43 | NA | NA | Caucasian |
| 6263 | T1D* | 31-35 | Male | 23.5 | 3.18 | 21 | NA | Hispanic/Latino |
| 6267 | AAb++ | 21-25 | Female | 23.5 | 16.59 | NA | GADA+<br>IA2A+ | Caucasian |
| 6271 | ND | 16-20 | Male | 24.4 | 11.47 | NA | NA | Caucasian |
| 6278 | ND | 11-15 | Female | 21.3 | 4.54 | NA | NA | African American |
| 6279 | ND | 16-20 | Male | 34 | 8.01 | NA | NA | Caucasian |
| 6282 | ND | 36-40 | Male | 41.9 | 6.83 | NA | NA | Caucasian |
| 6289 | ND | 16-20 | Male | 38.3 | 8.05 | NA | NA | African American |
| 6303 | AAb+ | 21-25 | Male | 31.9 | 3.03 | NA | GADA+ | Caucasian |
| 6310 | AAb+ | 26-30 | Female | 22.4 | 10.54 | NA | GADA+ | Hispanic/Latino |
| 6314 | AAb+ | 21-25 | Male | 23.8 | 1.49 | NA | GADA+ | Caucasian |
| 6318 | ND | 6-10 | Female | 17.6 | 3.89 | NA | NA | Caucasian |
| 6325 | T1D | 16-20 | Female | 31.2 | 0.14 | 6 | GADA+<br>IA2A+<br>mIAA+ | African American |
| 6335 | ND | 16-20 | Male | 23.6 | 8.85 | NA | NA | Multiracial |
| 6337 | T1D | 16-20 | Female | 17.9 | nd | 5 | mIAA+ | Caucasian |
| 6339 | ND | 21-25 | Male | 25 | 10.56 | NA | NA | Caucasian |
| 6362 | T1D | 21-25 | Male | 28.5 | 0.38 | 0 | GADA+ | Caucasian |
| 6366 | ND | 21-25 | Female | 20.5 | 0.41 | NA | NA | Hispanic/Latino |
| 6367 | T1D | 21-25 | Male | 25.7 | 0.39 | 2 | Negative | Caucasian |
| 6371 | T1D | 11-15 | Female | 16.6 | 0.11 | 2 | GADA+<br>IA2A+<br>mIAA+<br>ZnT8A+ | Caucasian |
| 6380 | T1D | 11-15 | Female | 14.6 | 0.22 | 0 | Negative | African American |
| 6384 | ND | 16-20 | Male | 18.2 | 0.7 | NA | NA | Caucasian |
| 6386 | ND | 11-15 | Male | 23.9 | 1.12 | NA | NA | Caucasian |
| 6388 | AAb++ | 21-25 | Female | 26 | 1.38 | NA | mIAA+<br>GADA+ | Hispanic/Latino |
| 6389 | ND | 16-20 | Male | 20.9 | 7.22 | NA | NA | Caucasian |
| 6396 | T1D | 16-20 | Female | 22.6 | 0.06 | 2 | Negative | Caucasian |
| 6397 | AAb+ | 21-25 | Female | 29.6 | 12.77 | NA | GADA+ | Caucasian |
| 6400 | AAb+ | 26-30 | Male | 22.2 | 4.17 | NA | GADA+ | Hispanic/Latino |
| 6401 | ND | 26-30 | Female | 31.3 | 12.81 | NA | NA | Hispanic/Latino |
| 6405 | T1D | 26-30 | Female | 42.5 | 1.84 | 0.6 | GADA+<br>IA2A+<br>ZnT8A+ | Hispanic/Latino |
| 6406 | ND | 6-10 | Male | 16.8 | 4.07 | NA | NA | Caucasian |
| 6424 | AAb++ | 16-20 | Male | 51.4 | 6.97 | NA | mIAA+<br>GADA+ | Hispanic/Latino |
| 6429 | AAb++ | 21-25 | Male | 19.6 | 2.25 | NA | mIAA+<br>GADA+ | African American |

**Supplementary Table 2.**
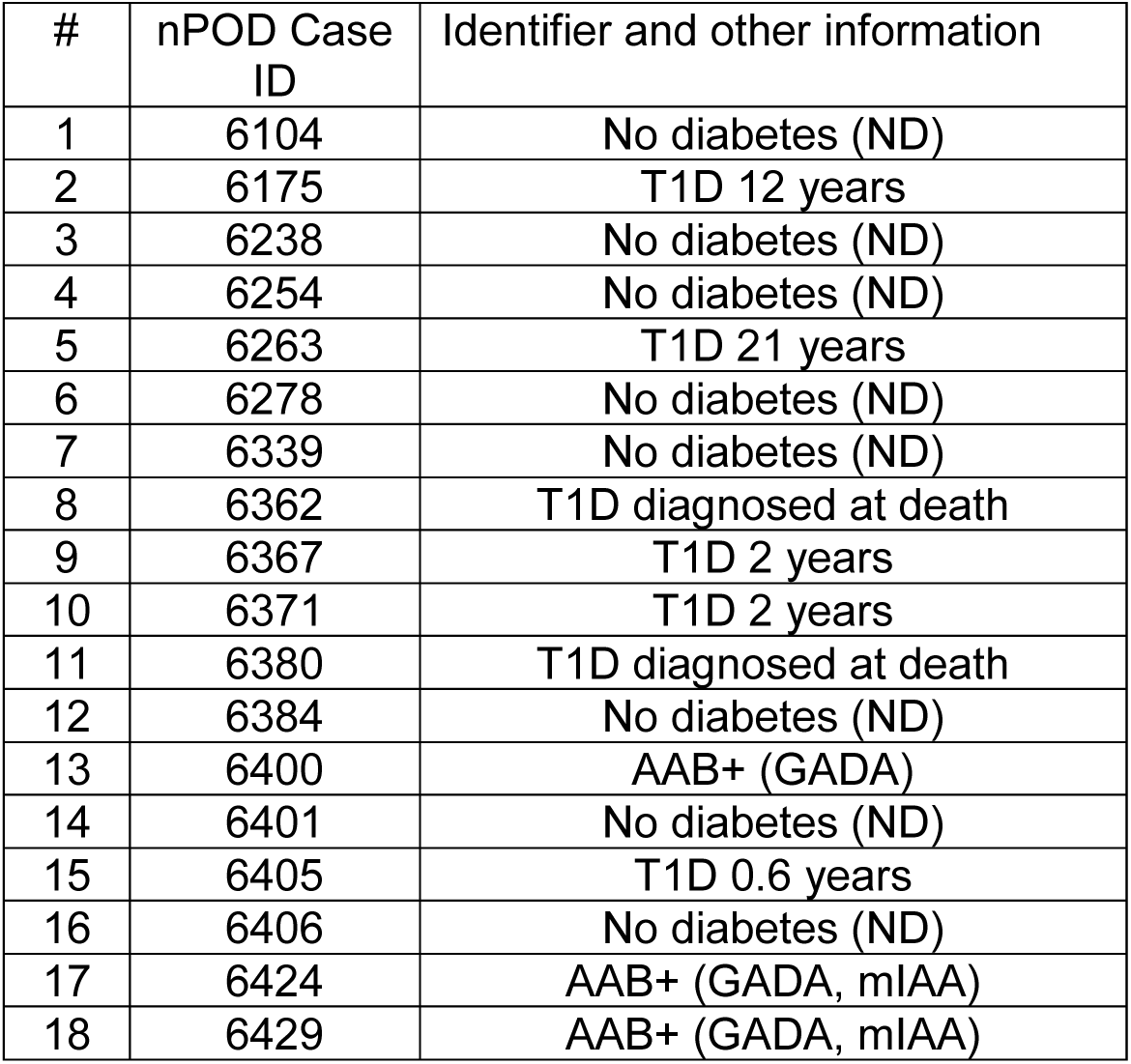
Donor phenotype and pancreas tissue samples. Additional information on these cases are already provided in supplementary Figure 1.

**Supplementary Table 3.** Age at passing, age of onset and disease duration among donors with type 1 diabetes with and without enterovirus infection by proteomics analysis.

| **Enterovirus negative** |  |  |  |
| --- | --- | --- | --- |
| Donor Number | Age at passing (years) | Age of Onset (years) | Disease duration (years) |
| 6046 | 16-20 | 6-10 | 8 |
| 6052 | 11-15 | 11-15 | 1 |
| 6070 | 21-25 | 11-15 | 7 |
| 6084 | 11-15 | 6-10 | 4 |
| 6195 | 16-20 | 11-15 | 5 |
| 6198 | 21-25 | 16-20 | 3 |
| 6245 | 21-25 | 11-15 | 7 |
| 6247 | 21-25 | 21-25 | 0.6 |
| 6325 | 16-20 | 11-15 | 6 |
| 6337 | 16-20 | 11-15 | 5 |
| 6362 | 21-25 | 21-25 | 0 |
| 6367 | 21-25 | 21-25 | 2 |
| **Mean** | **20.37** | **16.32** | **4.05** |
| **SD** | **3.92** | **4.97** | **2.72** |
| **Enterovirus Positive** |  |  |  |
| Donor Number | Age at passing (years) | Age of Onset (years) | Disease duration (years) |
| 6088 | 31-35 | 26-30 | 5 |
| 6113 | 11-15 | 11-15 | 1.58 |
| 6128 | 31-35 | 0-5 | 31.5 |
| 6143 | 31-35 | 26-30 | 7 |
| 6180 | 26-30 | 16-20 | 11 |
| 6209 | 0-5 | 0-5 | 0.25 |
| 6211 | 21-25 | 16-20 | 4 |
| 6212 | 16-20 | 11-15 | 5 |
| 6228 | 11-15 | 11-15 | 0 |
| 6371 | 11-15 | 6-10 | 2 |
| 6380 | 11-15 | 11-15 | 0 |
| 6396 | 16-20 | 11-15 | 2 |
| 6405 | 26-30 | 26-30 | 0.6 |
| **Mean** | **20.78** | **15.40** | **5.38** |
| **SD** | **9.46** | **7.95** | **8.48** |
|  | **P value** | **P value** | **P value** |
| **2-Tailed T-test** | **0.890** | **0.735** | **0.609** |

## Notes

### Competing Interest Statement

The authors have declared no competing interest.

### Author Declarations

University of Florida Health Center Institutional Review Board (IRB Protocol #201600029) The study of de-identified organ donor tissues is considered not to involve human subjects and is exempted from review by the Eastern Virginia Medical School Institutional Review Board (IRB)

